# Recommended Interventions for Enhanced Recovery After Cesarean Delivery in the United States

**DOI:** 10.1101/2024.11.05.24316713

**Authors:** Pervez Sultan, M Zakowski, Kareem Joudi, DJ Singh, Kimberly D. Gregory, D Lyell, Brendan Carvalho, Study investigators

## Abstract

**Background:** Existing enhanced recovery after cesarean delivery (ERAC) professional society recommendations lack intersociety collaboration, multidisciplinary expert input and involvement of patient stakeholders. This initiative aimed to develop a multidisciplinary set of ERAC interventions endorsed by relevant US professional societies and patient representatives.

**Methods:** This American Society of Anesthesiologists (ASA) and American College of Obstetricians and Gynecologists (ACOG) led initiative received IRB approval from Stanford University. In total 19 stakeholders were invited to participate in this Delphi study, including 13 representatives from 8 US professional societies (American Society of Anesthesiologists (ASA - 1 representative), Society for Obstetric Anesthesia and Perinatology (SOAP - 2), American College of Obstetricians and Gynecologists (ACOG - 4), Society for Maternal Fetal Medicine (SMFM - 1), Society of OBGYN Hospitalists (SOGH - 1), Enhanced Recovery After Surgery Society (ERAS Society - 1), Association of Women’s Health Obstetric and Neonatal Nurses ((AWHONN -2), American Association of Nurse Anesthesiology (AANA - 1), a physical therapist, a lactation expert and 4 patient representatives from diverse backgrounds. A three-round modified Delphi approach was conducted (2 rounds of electronic questionnaires and a 3^rd^round of e-discussion), to produce the final set of ERAC recommendations. An initial list of 70 interventions was compiled based on a previously published systematic review of ERAC studies and professional society guidelines/recommendations (ERAS Society, SOAP, and Healthcare Canada). Consensus was obtained for the final list of interventions, with strong consensus defined as ≥70% agreement and weak consensus as 50–69% agreement.

**Results:** All professional societies that were approached provided content-expert stakeholders to participate, and all stakeholders completed all rounds of the Delphi process. A final set of 32 interventions (6 preoperative, 13 intraoperative and 13 postoperative) achieved strong consensus.

**Conclusions:** This US intersociety and multidisciplinary Delphi study resulted in a list of ERAC interventions, which should be considered when implementing ERAC within US labor and delivery units.

## Introduction

Maternal morbidity and mortality in the United States is rising and improving maternal health is one of the World Health Organization’s (WHO) key priorities.^1^ Enhanced Recovery After Surgery (ERAS) protocols are evidence-based interventions developed by multidisciplinary teams and key stakeholders. ERAS protocols are proven to be an effective strategy to optimize patient experience and satisfaction, while reducing morbidity and healthcare expenditure in multiple surgical specialties including colorectal,^2–6^ vascular,^7^ thoracic^8^ and urological surgery.^9,10^ Cesarean delivery is the most performed inpatient operation worldwide, with over 1 million surgeries performed annually in the US alone.

High quality, simple interventions, before, during and after cesarean delivery combined with system changes can improve maternal and neonatal outcomes. ERAS programs have demonstrated success in obstetric settings,^11–13^ significantly reducing length of hospital stay, improving pain outcomes, reducing hospitalization costs,^14–16^ and complication rates^15,17^ without increasing maternal readmssion. The Society of Obstetric Anesthesia and Perinatology (SOAP) states that enhanced recovery for cesarean delivery (ERAC) is an essential criteria for Centers of Excellence designation.^18^ Several professional society guidelines support the principles of enhanced recovery in the cesarean delivery setting including publications by the: ERAS Society,^19–21^ SOAP,^22^ American College of Obstetricians and Gynecologists (ACOG),^23^ National Institute for Health and Care Excellence (NICE)^24^ and Healthcare Excellence Canada.^25^ However, current ERAC interventions vary across published guidelines, lack inter-society collaboration, US multidisciplinary expert input and involvement of patient stakeholders.

The success of ERAC is attributed to a combination of protocol interventions, implementation strategies, monitoring practices and evaluation tools.^26^ Recommendations for cesarean delivery interventions to improve maternal and neonatal care and outcomes require revision and review based on contemporary evidence and multidisciplinary expert opinion including academic and rural clinicians, obstetricians, MFMs, anesthesia providers, nursing, midwives, lactation specialists, physical therapists and neonatologists. The development of contextually relevant guidelines that confer clinical benefit for patients undergoing cesarean delivery in the US would provide a starting point to standardize best peripartum care, and form the basis of best ERAC practices.

The aim of this Delphi study led by the American Society of Anesthesiologists (ASA) and ACOG was to develop US specific ERAC intervention recommendations through expert stakehodler input and endorsement from US multidisciplinary professional societies.

## Methods

Following institutional review board approval from Stanford University (ID number 70012) we conducted a 3 round Delphi study to develop US multidisciplinary recommendations for ERAC. These recommendations were designed specifically for patients undergoing scheduled cesarean delivery in the US.

### Phase 1. Identify and approach professional society stakeholders and patient representatives

The ASA committee for obstetric anesthesia approved a proposal to lead this initiative in collaboration with the ACOG. US professional multidisciplinary societies were then approached to provide expert stakeholders for inclusion in the planned Delphi study. Professional societies were approached and asked to nominate candidates based on their specific expertise and interest in ERAC. In total 19 stakeholders were invited to participate in this Delphi study, including 13 representatives from 8 US professional societies: ASA - 1 representative, SOAP - 2, ACOG - 4, Society for Maternal Fetal Medicine (SMFM - 1), Society of OBGYN Hospitalists (SOGH - 1), ERAS Society - 1, Association of Women’s Health Obstetric and Neonatal Nurses (AWHONN -2), American Association of Nurse Anesthesiology (AANA - 1), a physical therapist, a lactation expert and 4 patient representatives. Patient representatives were invited who had experienced cesarean delivery resulting in a live birth in the US within the previous 5 years.

### Phase 2. Identify ERAC interventions

We generated a comprehensive list of ERAC interventions from existing published guidelines/ recommendations/consensus statements (ERAS Society, SOAP, and Healthcare Canada^19–25^) and from published ERAC versus control studies identified in a previous systematic review and 2 published meta-analyses.^11–13^ Following removal of duplicates, a list of 70 interventions were compiled for consideration in Round 1 of the Delphi process. A full list of these interventions divided into pre-, intra- and postoperative phases of care is provided in Appendix 1.

### Phase 3. Delphi consensus surrounding items to include in multidisciplinary recommendations

The Delphi process involves an iterative process of multiple rounds, including generation of long lists (selection of existing interventions and generation of new interventions where required), feedback and voting.^27–30^ Modified Delphi methodology includes at least 2 rounds of electronic questionnaires followed by a 3^rd^ round for round-table discussion and ratification.^29^ This study followed a modified Delphi approach with 2 rounds of electronic questionnaires followed by a round table discussion. The study was conducted by an Executive Committee (PS, SJ and BC) who conceived, designed and executed the study with a panel of 19 stakeholders.

The aim of the Delphi consensus was to produce a set of recommended multidisciplinary ERAC interventions for patients undergoing scheduled cesarean delivery in the US setting.

#### Round 1 (August 2023)

70 Interventions identified from existing guidelines, recommendations and ERAC versus control studies were screened by expert stakeholders to determine the most applicable or appropriate interventions for use in US labor and delivery units. Stakeholders were invited to score each intervention between 1-9 (1-3=intervention is ‘of limited importance or invalid,’ 4-6=intervention is ‘important but not critical for inclusion or requires revision’ and 7-9= intervention is ‘critical for inclusion’),^31–33^ as recommended by the Grading of Recommendations Assessment, Development and Evaluation (GRADE) working group for assessing research evidence level of importance.^34,35^ Proposed details surrounding the interventions were planned to be finalized in round 3 if selected. Stakeholders were invited to amend or edit as appropriate using free text. Pre-defined drop-down menus were used wherever possible with space to provide alternatives and general feedback. Responses were returned to the project administrator (KJ) for anonymization and collation. A fully anonymized spreadsheet containing all comments and score selections were subsequently analyzed by the Executive Committee and revisions were made as required for Round 2 along with explicit justification for any changes made. For an outcome to proceed to Round 2 it required: (i) a score of ≥7 selected by ≥70% of stakeholders, or (ii) a score between 4-6 selected by ≥70% of stakeholders (i.e. the number of ≥7 scores and 4-6 scores were not combined, and strong consensus was required to proceed to the next round). Outcomes were excluded if they did not meet the above criteria or if a score between 1-3 was selected by ≥70% or stakeholders. A cut-off value of 70% was selected by the Executive Committee following review of the COMET Handbook.^36^ No broad agreement exists regarding what constitutes consensus,^37^ however, this approach is supported by a previously published study evaluating a long list of outcomes similar to that proposed in this study.^38^

#### Round 2

In October 2023 all expert stakeholders that participated in Round 1 received an anonymized summary of the results, including numbers of selections for scores between 1-3, 4-6 and 7-9 and median scores (with 25^th^ and 75^th^ percentiles) for each outcome. In Round 2, stakeholders were invited to score each outcome that met the criteria to proceed to Round 2. Interventions were once again scored between 1-9 utilizing dropdown menus. Free text alternatives were encouraged if stakeholders felt that an appropriate option was not included. These would be considered in the Round 3 discussion. The criteria for an outcome to proceed to Round 3 were the same as required to proceed from Round 1 to Round 2.

Patient representatives were invited to rank 14 ERAC interventions based on their perceived importance from their perspective (ranking of 1-14 where 1 is most important and 14 is least important). These interventions were based on themes identified from existing ERAC literature and guidelines. An undesirable intervention (‘intervention that worsens recovery’) was also included in the list to verify patient representative understanding of the task. The responses would only be analyzed if the undesirable intervention was ranked as 14. The findings from the patient representatives were provided to all participants of the Round 3 discussion.

#### Round 3

Based on optimal availability, stakeholders were invited to attend a recorded electronic round-table discussion on December 13^th^ 2023, (using Zoom Video Communications software, San Jose, California), aiming to finalize and achieve consensus on the multidisciplinary ERAC interventions. The session was chaired by an Executive Committee member (PS) and 1 co-chair (BC). Comments and median scores from Round 1 and 2 were included and excluded interventions were disclosed to all stakeholders prior to the meeting. Interventions which met the criteria for inclusion in Round 3, were discussed among available stakeholders. Discussion was limited to 5-minutes per intervention and guided by the following factors: 1) Evidence supporting potential benefits of the intervention; 2) Current utilization in clinical practice; 3) Feasibility: practicality or ease of implementation; and 4) Cost of intervention. Following discussion, stakeholders were invited to participate in live anonymized online polling, with stakeholders voting to either ‘include,’ ‘exclude’ or ‘do not feel qualified to respond’ for each proposed intervention. Using an iterative process, areas that warranted revision were modified and subsequent voting was undertaken.

Once interventions that met Round 3 criteria were discussed and voted upon, the stakeholders were given the opportunity to: 1) re-introduce previously excluded interventions from Rounds 1 or 2, and 2) introduce new interventions that had not previously been used in the published ERAC literature, and therefore had not been considered thus far in the Delphi process. Any outcome that was re-introduced or introduced for the first-time during Round 3 required a stakeholder to propose the outcome, a separate participant to second it and provide a brief justification to the remaining Delphi stakeholders, prior to it being considered and voted upon. The Round 3 virtual round table discussion was limited to 2 hours (13^th^ December 2023) and therefore discussion was continued on January 23^rd^ 2024, in order to complete the proposed Round 3 tasks. In Round 3, interventions were classified as a proportion of present stakeholders as follows:^36,39^

1. ≥ 70% agreement among those that felt qualified to vote = strong consensus; intervention included in the final core intervention list.
2. 50–69% agreement among those that felt qualified to vote = weak consensus; include in manuscript Table legend acknowledge as an intervention to consider when designing an ERAC clinical protocol.
3. < 50% agreement among those that felt qualified to vote = intervention excluded as a core intervention.

Digital recordings of the Round 3 discussions were sent to all stakeholders who completed Rounds 1 and 2 but could not attend Round 3. All stakeholders were asked to vote using drop down menus, after listening to the Zoom recording of the Round 3 discussion.

Upon finalization of the developed recommendations, all stakeholders and professional society representatives were invited to approve the finalized recommendations prior to publication.

### Statistical analysis

A minimum of 17 stakeholders was the desired target in this study; the median [interquartile range] and range of participants in Delphi studies have previously been reported as 17 [11-31] and 3 to 418, respectively.^37^ Data were reported descriptively. Spreadsheets were developed for each round and circulated in Microsoft Excel (Excel for Mac; version 16.49, 2021) spreadsheet format. All denominator values for percentages were based on responses, and percentage values reported signify the proportion of stakeholders in agreement with a particular option.

## Results

All professional societies that were approached provided content-expert stakeholders to participate in the Delphi process. A summary of expert specialty, US state of practice and area of expertise is provided in Appendix 1 and a summary of stakeholder demographic details is provided in Appendix 2. All stakeholders completed all rounds of the Delphi process, with live attendance of the round 3 virtual meetings by 12 and 8 stakeholders, respectively.

### Rounds 1 and 2

A full list of interventions considered in Round 1 divided into pre, intra and postoperative is provided in Appendix 3. In total, 70 interventions were considered in Round 1 (16 preoperative, 26 intraoperative, 28 postoperative), and 66 interventions in Round 2 (13 preoperative, 26 intraoperative, 27 postoperative).

#### Round 3

43 interventions were discussed and voted upon in round 3 (10 preoperative, 15 intraoperative, 18 postoperative). Among these included interventions were 2 interventions that were re-introduced (promotion of rest periods, and breastfeeding support and education).

#### Final list of recommended recommendations

A final set of 32 interventions (6 preoperative, 13 intraoperative and 13 postoperative) achieved strong consensus for recommendation. The interventions which achieved a strong consensus during the Round 3 discussion (≥70% votes to include), proposed descriptions or examples to provide clarity to these interventions where applicable, and proportion of votes are provided in Table 1.

**Table 1:**
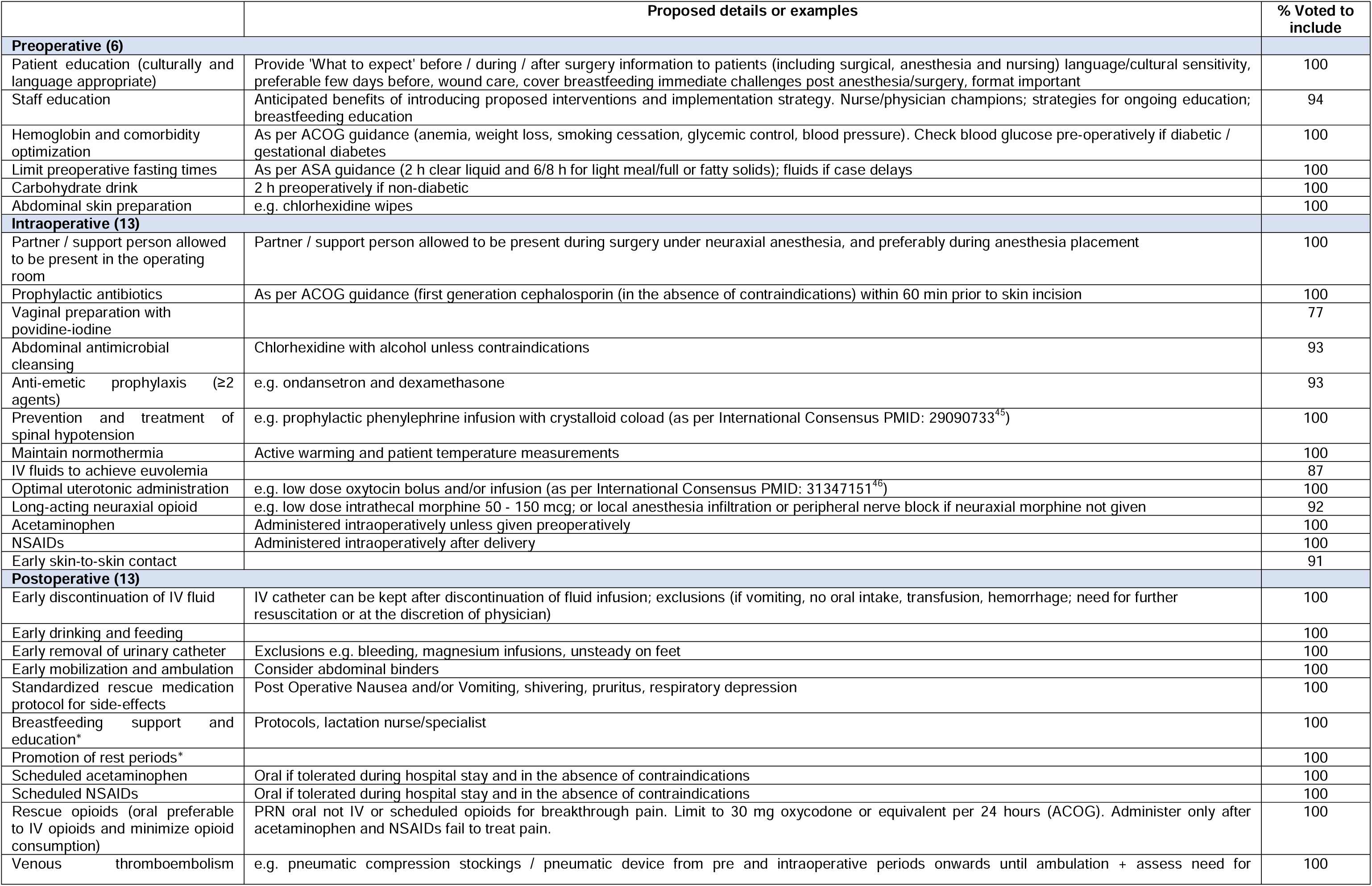
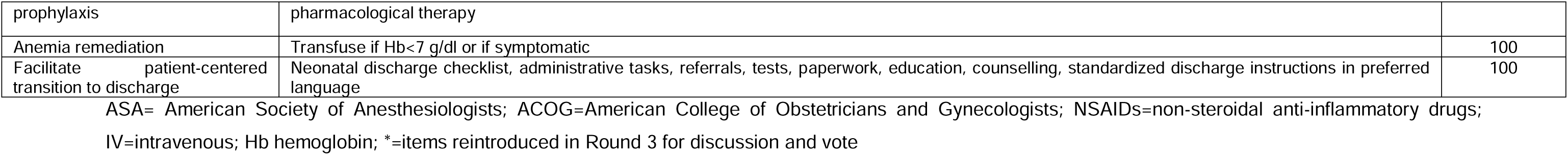
Recommendations receiving strong consensus among US multidisciplinary experts and patient representatives.^45,46^.

One intervention considered during the Round 3 discussion (preoperative antacid prophylaxis) achieved weak consensus. This intervention received 59% (i.e. between 50-69%) of votes and therefore was not included in the final set of recommendations.

## Discussion

### Main findings

We have developed a set of ERAC recommendations using a Delphi process and involving a diverse range of multidisciplinary stakeholders that included representatives from all major US professional societies relevant to caring for patients undergoing cesarean delivery. These interventions should be considered when implementing or refining an ERAC protocol for patients undergoing cesarean delivery.

### Clinical implications

Enhanced recovery after surgery principles include improving quality of care and outcomes, optimizing patient experience and reducing cost through the implementation of evidence-based recommendations.^40,41^ These recommended interventions were derived from a comprehensive review of contemporary literature, which were then selected by relevant stakeholders.

Stakeholders were selected from all major US professional societies, which treat birthing patients and included patient representatives from underrepresented minority groups. These recommended interventions by stakeholders with either significant clinical expertise and or lived experience of cesarean delivery can be considered best practice interventions for ERAC protocols. Implementation of these recommended interventions could help to reduce variability in practice among institutions in the US, and ultimately improve outcomes following this commonly performed inpatient surgical procedure.

Existing ERAC recommendations (ERAS Society, SOAP, ACOG and Healthcare Excellence ^19– 23,25^) were not developed through a Delphi process, and were developed without adequate transdisciplinary representation of professional societies. Furthermore, patient representatives were not included in previous ERAC recommendations. Patient representatives are important for Delphi processes and guideline development.^36^ We included patients from minority groups and diverse backgrounds in our stakeholder group discussions. The recommended interventions selected were based on available evidence and with consideration of feasibility for implementation given current healthcare systems and processes within US delivery units.

While many of the recommended interventions have been included in previously published statements and recommendations, several important differences are worthy of note. Firstly, presence of a partner or relative during surgery was felt to be important among stakeholders as a strategy to reduce anxiety and provide important support to the patient during surgery.

Metoclopramide prophylaxis was also not included in the recommendations as the number needed to treat to prevent aspiration pneumonitis in patients undergoing cesarean delivery which are mostly performed with neuraxial anesthesia, was felt to be too high compared to the risk of undesirable side-effects such as extrapyramidal symptoms.

### Research implications

While this initiative has recommended interventions to consider as part of ERAC protocol, the best implementation strategy is yet to be determined. There appears to be large heterogeneity among published ERAC versus control studies regarding implementation strategies used including multidisciplinary stakeholder involvement, process mapping, duration of implementation phase, data collection following implementation and how to best demonstrate sustained change in practice.^42^ Furthermore, there appears to be inconsistent use of SQUIRE (Standards for Quality Improvement Reporting Excellence) guidelines among these published studies.^11^

Despite numerous studies evaluating ERAC versus control group, the GRADE (Grading quality of evidence and strength of recommendations) levels of evidence supporting ERAC remain either low or very low for most outcomes, largely due to the lack of high-quality studies in this field.^11,13^ Most studies exploring the impact of ERAC have utilized an observational study cohort design. This may be the optimal study design as it facilitates system and cultural change amongst multidisciplinary healthcare workers. Only 3 randomized controlled trials have been reported in previous reviews, which consequently limits the availability of high quality evidence in this field.^16,17,43^ The lack of randomized controlled trials is compounded by the high heterogeneity among studies in terms of interventions and outcomes that have been used. US centers considering implementing ERAC interventions can evaluate its impact using 15 core outcome measures that were recommended in a Delphi consensus study involving 32 international experts and patient representatives.^44^ These impact metrics together with our newly developed multidisciplinary recommendations could help to improve levels of evidence supporting ERAC in future studies. The use of cluster randomization study design (where entire hospitals are randomized to implement ERAC), step-wise implementation or multicenter implementation studies may also improve levels of evidence supporting the use of ERAC.

### Strengths and limitations

The main strengths of this study are the utilization of a formal modified Delphi study design, which has not previously been included as part of published ERAC recommendations.

Furthermore, the inclusion of multidisciplinary stakeholders from all major US professional societies identified as most relevant to ERAC, as well as a diverse group of patient representatives, provided broad representation to this process.

We do however acknowledge some limitations. We focused on interventions that are most relevant to the US healthcare setting. Therefore, certain drugs that are not available in the US were not included in these recommendations for example, carbetocin (a uterotonic agent) and diamorphine (a long-acting opioid). Similarly, some of the recommended interventions may not be feasible in low resource settings where maternal mortality rates are known to be highest and where patients may benefit most from a country specific ERAC protocol. For example, the lack of pharmacy staff and drug availability may preclude the administration of phenylephrine infusions to prevent spinal hypotension, equipment availability may preclude intraoperative patient warming, nurse staffing ratios may be low thus reducing proportion of patients achieving early ambulation and urinary catheter removal targets. We encourage the development of guidelines in other countries to ensure that feasible and appropriate interventions are recommended for implementation by healthcare providers within the context of their own clinical practice. Our recommendations could be a starting point for context-specific ERAC protocol development.

We also acknowledge that cesarean delivery accounts for only 20% of global births (approximately 30% of US births), and the recommendations we provide are specific to scheduled cesarean delivery. Therefore, specific recommendations for enhanced recovery after intrapartum cesarean delivery, operative vaginal delivery, and spontaneous vaginal delivery, which constitute most global births, are still needed. Standardization of care pathways may ultimately help to identify and address disparities in cesarean delivery care. Beyond delivery mode, enhanced recovery guidelines developed for populations that are known to be at higher risk for postpartum complications should be considered for different racial and ethnic groups, patients that experience severe maternal morbidity and patients who screen positive for psychological morbidity.

### Conclusion

In summary, we have developed a set of ERAC recommendations from a diverse range of multidisciplinary stakeholders, including patients, and all major US professional societies relevant to peripartum patients. These interventions should be considered for implementation in US labor and delivery settings for patients undergoing cesarean delivery.

## Data Availability

All data produced in the present work are contained in the manuscript

