## Appendix 1 for "Recommended Interventions for Enhanced Recovery After Cesarean Delivery in the United States"

**Appendix 1. List of Expert Stakeholders Participating in the Development of the Intervention Recommendations for Enhanced Recovery After Cesarean delivery (ERAC)**

| **Expert name** | **Society or specialty represented and place of work** |
| --- | --- |
| **American College of Obstetricians and Gynecologists (ACOG)** | |
| Brook Thomson | Associate Professor, Associate Medical Director for inpatient obstetrics, University of Texas Health Science Center at San Antonio, TX |
| Gabriela Dellapiana | Staff Physician, OB GYN – Maternal Fetal Medicine, Cedars-Sinai, LA, CA |
| Kim Gregory | Professor, Director of Maternal-Fetal medicine, Vice Chair of Women’s Health care quality and performance improvement, Cedars-Sinai, LA, CA |
| Deepjot Singh | Co-Chair of ACOG Committee Patient safety and quality improvement, Chair of Medical Staff Performance Improvement Torrance Memorial/Cedars-Sinai affiliate, Providence, Torrance, CA |
| **American Society of Anesthesiologists (ASA)** | |
| Mark Zakaowski | Professor, Fellowship Director of Obstetric Anesthesiology, Cedars-Sinai, LA  ASA Obstetric anesthesia subcommittee Chair  Co-author on SOAP consensus statement on ERAC |
| **Society for Obstetric Anesthesia & Perinatology (SOAP)** | |
| Pervez Sultan | Associate Professor, Stanford University School of Medicine, Palo Alto, CA. Co-author on SOAP consensus statement on ERAC |
| Brendan Carvalho | Professor, Stanford University School of Medicine, Palo Alto, CA  Co-author on SOAP consensus statement on ERAC |
| **Society for Maternal Fetal Medicine (SMFM)** | |
| Mara Greenberg | Kaiser Permanente, Oakland Medical Center, Oakland, CA |
| **Enhanced Recovery After Surgery (ERAS) Society** | |
| Gregg Nelson | Deputy head of Obstetrics and Gynecology, Cumming School of Medicine, Calgary, Alberta, Canada  ERAS Society Treasurer Executive Committee  Co-author of ERAS Society guidelines for cesarean delivery and women’s health chapter |
| **Society of OBGYN Hospitalists (****SOGH)** | |
| Rakhi Dimino | Medical Director of Operations at Ob Hospitalist Group, Houston, TX |
| **Association of Women's Health, Obstetric and Neonatal Nurses (AWHONN)** | |
| Jessica Irrobali | AWHONN Director of Collaborative Clinical Programs.  St. David’s Women’s Center of Texas, Austin, TX |
| Cheryl Parker | Assistant Professor, Director for the Doctor of Nursing Practice, Nurse Anesthesia track at University of Louisville, KY  AWHONN Kentucky State Chair and Alliance for Innovation on Maternal Health (AIM) representative |
| **American Association of Nurse Anesthesiology (AANA)** | |
| Brett Morgan | AANA Sr Director, Education and Practice  Western Carolina University, Durham, NC |
| **Other experts: Physical therapist representative** | |
| [Lauren](https://www.linkedin.com/in/lauren) Jarmusz | Department of physical therapy, Stanford University, Palo Alto, CA |
| **Other experts: Lactation expert** | |
| Allison Walsh | International Board-Certified Lactation Consultant and doula, New York |
