## Appendix 2 for "Recommended Interventions for Enhanced Recovery After Cesarean Delivery in the United States"

**Appendix 2.** **Summary of stakeholder demographics**

|  | **Stakeholders**  **(n=15)** | **Patient representatives**  **(n=4)** |
| --- | --- | --- |
| Race  White  Asian  Black  Other | 10  3  2  - | 2  0  2  - |
| Ethnicity  Non-Hispanic  Hispanic | 14  1 | 1  3 |
