## Appendix 3 for "Recommended Interventions for Enhanced Recovery After Cesarean Delivery in the United States"

**Appendix 3. List of ERAC interventions considered in Round 1**

**Preoperative interventions (16)**

Patient education

Patient counselling - lifestyle

Staff education

Breastfeeding preparation and information

Weight management, blood pressure, glycemic control if applicable

Hemoglobin optimization

Preoperative identification of patients appropriate for enhanced recovery

Midwife consultation

Limit preoperative fasting times

Antacid prophylaxis

Optimization of gastric emptying

Carbohydrate drink

Abdominal skin preparation

Check blood glucose pre-operatively if diabetic / gestational diabetes

Acetaminophen administered preoperatively

NSAID administered preoperatively

NSAID=non-steroidal anti-inflammatory drug

**Intraoperative interventions (26)**

Prophylactic antibiotics

Maintain normothermia

IV fluids to achieve euvolemia

Fluid restrictive regimen

Partner / support person allowed to be present during surgery under neuraxial anesthesia

Long-acting neuraxial opioid

Prevention of spinal hypotension

Treatment of spinal hypotension

Anti-emetic prophylaxis (single agent)

Anti-emetic prophylaxis (≥2 agents)

Peripheral nerve/fascial plane blocks

Local anesthesia wound infiltration (single-shot)

Local anesthesia wound infiltration (continuous infusion)

Acetaminophen administered intraoperatively

NSAIDs administered intraoperatively

Optimal uterotonics

Delayed cord clamping

Early skin to skin contact

Vaginal preparation with povidine-iodine

Abdominal antimicrobial cleansing

Sub-cuticular wound closure

Closure of hysterotomy in 2 layers and non-closure of peritoneum

If >2cm subcutaneous tissue, re-approximation of tissue layer

Blunt expansion of transverse uterine hysterotomy

Air not oxygen for neonatal resuscitation

Maintain normothermia

NSAIDs=non-steroidal anti-inflammatory drugs

**Postoperative interventions (28)**

Early mobilization and ambulation

Early removal of urinary catheter

Early drinking

Early feeding

Sham feeding

Day 1 blood tests

Anemia remediation

Venous thromboembolism prophylaxis

Scheduled course of antibiotics

Early dressing removal

Incentive spirometry

Routine glucose monitoring, and tight control if treated

Standardized rescue medication protocol for side-effects

Early removal of IV fluid

Early removal of IV cannula

Acetaminophen

NSAIDs

IV acetaminophen and/or NSAIDs for prescribed period

Oral opioids

Minimize opioid consumption

Breastfeeding support / education

Infant temperature monitoring

Neonatal discharge checklist

Early communication with community midwife or carer

Early maternal discharge

Discharge education and counselling

Promote resting periods

Standardized written discharge instructions

IV=intravenous; NSAIDs=non-steroidal anti-inflammatory drugs
